# Monitoring Heart Failure Patients with Mobile Health Technologies: Outcomes from a 180-day Prospective Study

**DOI:** 10.1101/2023.08.30.23294869

**Authors:** Sukanya Mohapatra, Mirna Issa, Vedrana Ivezic, Rose Doherty, Stephanie Marks, Esther Lan, Keith Rozett, Lauren Cullen, Wren Reynolds, Rose Rocchio, Gregg C. Fonarow, Michael K. Ong, William F. Speier, Corey W. Arnold

## Abstract

Mobile health (mHealth) methods have grown in popularity in preventative medicine due to their convenience, cost, and ability to acquire actionable data. At the same time, the burden of many diseases has grown due to their prevalence and high rates of morbidity and mortality. To combat this burden, mHealth can be customized to improve adherence to recommended regimens and decrease doctor or hospital visits for patients and their families. Heart failure (HF) is a disease with an especially high burden for which mHealth can be used to promote adherence to advised care plans with the goal of decreasing exacerbations and their associated urgent and emergency care visits. Our study compared adherence to different mHealth monitoring regimens that used activity trackers, scales, and a mobile app with gamification features and a financial incentive. In a prospective analysis of 111 HF patients monitored for 180 days, we found that a regimen including a mobile app with a gamified financial incentive led to significantly higher adherence to activity tracker (95% vs. 72.2%, p=0.0101) and weight (87.5% vs. 69.4%, p=0.0016) monitoring compared to a regimen that included the monitoring devices alone. Our findings indicate that mobile apps with added engagement features can be useful tools to reduce temporal adherence decline and may thus increase the impact of mHealth driven interventions.

## Introduction

Heart failure (HF) is a severe health condition associated with high morbidity, mortality, and health resource use (Al Olama et al., 2015). In the United States, 6.2 million adults have HF, and roughly $31 billion is spent annually on HF-related costs (Virani et al., 2020). Approximately 400,000 death certificates in 2018 cited HF and it is predicted that by 2030, cases will surge by 25% (Nelson, 2021; Benjamin et al. 2019; Virani et al., 2020). The relatively high prevalence of this chronic disease and associated morbidity and mortality impose a substantial economic burden on healthcare expenditures. An alarming cost increase is projected, with a total of up to $69.7 billion by 2030, a 127% growth since 2012 (Virani et al., 2020).

The primary contributor to HF spending is hospitalization, accounting for 75-80% of total costs (Heidenreich et al., 2013). With approximately one million annual hospitalizations, HF is a leading cause of hospital admissions in people over 65 (Mcdermott et al., 2005). Readmissions among HF patients are, in particular, a significant concern. Approximately 1 in 4 patients are readmitted within 30 days of discharge, and 1 in 2 patients are readmitted within six months of discharge (Nelson et al., 2021). Clinical guidelines suggest that adherence to self-care recommendations such as regular medication intake, weight monitoring, and physical activity is vital to a favorable prognosis (Lainscak et al., 2011; Ponikowski et al., 2016; Riegel et al., 2009). Nearly half of all HF readmissions are preventable with enhanced adherence to such self-care behavior after discharge (Michalsen et al., 1998; Tsuyuki et al., 2001; Windham et al.). There is a clear need to identify effective adherence-enhancing tools and methods to promote positive HF health outcomes.

Mobile health (mHealth), using mobile interventions to support health, may be a preferable and less intensive method to deliver medical care (Hale et al., 2015; Kumar et al., 2013; Steinhubl et al., 2015). By including the use of mHealth technologies in the home monitoring of HF, patients may be more inclined to play an active role in lifestyle modifications that are intended to improve their health outcomes and prevent rehospitalization (Feldman et al., 2018). The use of mobile applications can supplement these mHealth interventions by providing patients with an avenue to monitor their progress and receive adherence notifications. Such apps are designed to serve as a helpful reminder for patients to complete their daily self-care tasks, potentially increasing the effectiveness of telemedicine interventions, including those involving mHealth technologies (Jaarsma et al., 2000).

The use of mHealth has been expanding to include more diverse patient populations (Fiordelli et al., 2013). Attributable to their remote nature and resulting accessibility, mHealth technologies have extended into smartphone applications, imaging services, and other technological functions. These developments can enhance support for patients by enabling them to complete health and symptom monitoring surveys, providing education on risk factors, and allowing patients to monitor and track health metrics continuously and in real time. Present-day home monitoring interventions employ wireless sensors, telephone services, websites, and home visits from nurses (Suh et al., 2011; Wakefield et al., 2009; Zan et al., 2015a). Several studies using mHealth in HF patients have suffered from small sample size and poor patient adherece to monitoring (Chaudhry et al., 2013; Gensini et al., 2017; Ong et al., 2016; Ware et al., 2019; Zan et al., 2015b). For instance, one trial found that only 55.4% of patients randomized to telemonitoring used the given technology-based intervention at least half of the time at month one, and 51.7% at month six (Gensini et al., 2017). The reported low adherence levels and inconclusive results associated with such studies are in part attributable to the high monitoring burden of home interventions. In investigating strategies to overcome these barriers, the combined use of gamification and financial incentives has yet to be extensively employed in mHealth adherence studies (Tran et al., 2022).

This study evaluated differences in adherence levels to three distinct mHealth home monitoring regimens in HF patients involving a combination of devices, a mobile app, and financial incentives with the hypothesis that including a mobile app would boost adherence to study devices. Device and app usage data from participants in each regimen were compared using statistical analyses to identify any differences. Our results provide new insights into tailoring mHealth interventions to drive adherence.

## Study Design/Methods

### Study Population

This prospective study included patients with heart failure at the University of California, Los Angeles (UCLA) medical center. The study was approved by the UCLA Institutional Review Board (IRB). Patients with an HF diagnosis were identified using our institution’s electronic health record (EHR). English-speaking adults aged 50-80 who had a diagnosis of HF and owned a smartphone were eligible to participate in the study. Exclusion criteria included participants who were receiving hemodialysis, had received an organ transplant or were on an organ transplant waiting list, or did not have the cognitive or physical ability to participate.

### Subject Recruitment

Figure 1 overviews our study population and recruitment procedures. Individuals who met the eligibility criteria were contacted by email and text to gauge interest in the study. Over the phone, research personnel contacted interested individuals to provide them with additional information and conducted the verbal consent process with those interested in participating. To ensure an interested individual was capable of using their smartphone, they first participated in a daily survey delivered via text messages for one week. The survey consisted of three questions: 1) how much did symptoms of heart failure limit your life yesterday? 2) were you able to take your medication as prescribed yesterday? and 3) did you follow your recommended diet yesterday? Participants who completed at least five of the seven surveys were invited to participate in the full study for 180 days.

**Figure 1.**
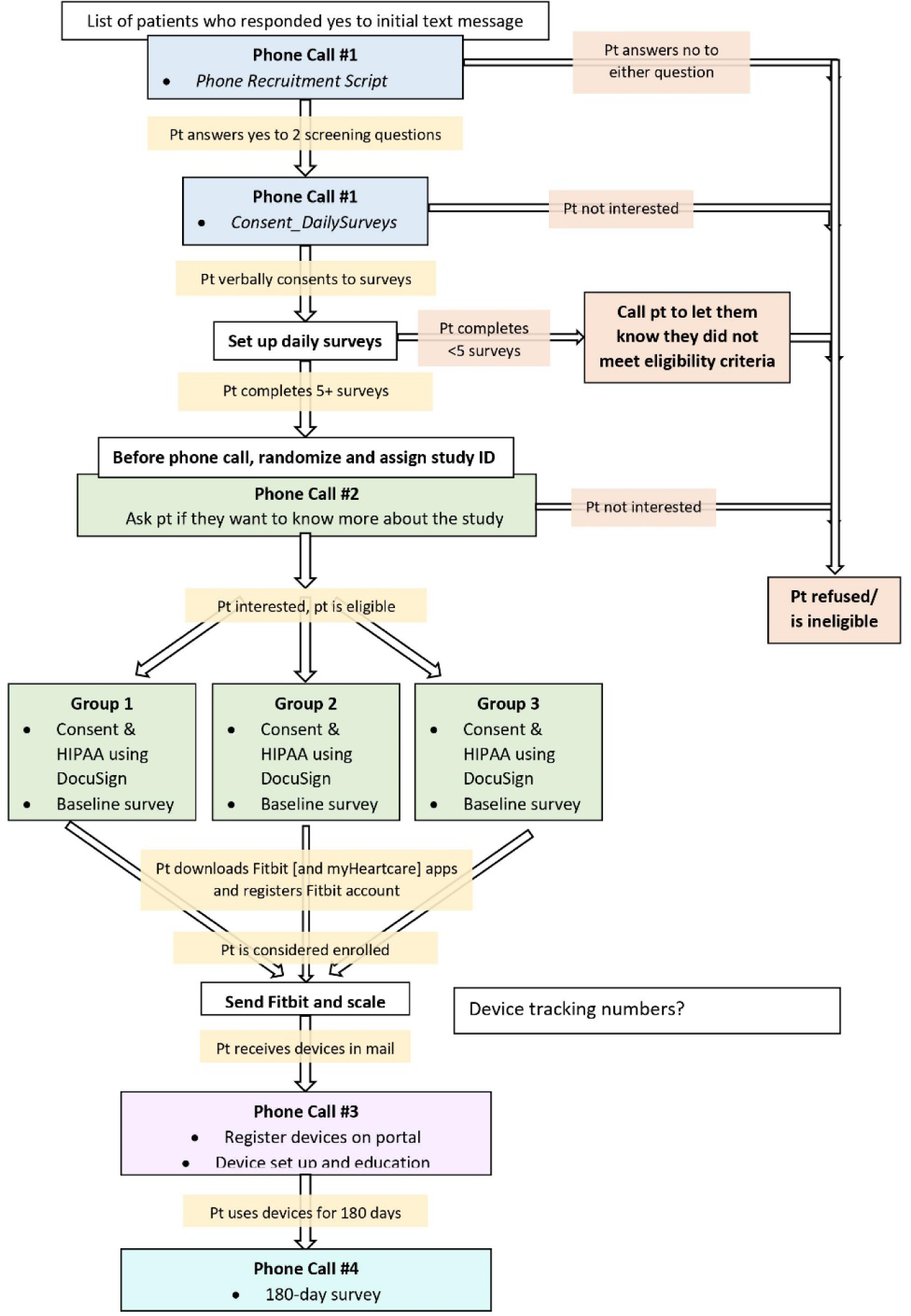
Enrollment flow diagram detailing the steps between recruiting patients with an initial message to eventually surveying responses at the conclusion of their study period.

### Study Groups

To investigate the impact of different mHealth regimens on adherence, three groups used various combinations of an activity tracker, a scale, a mobile app, and a financial incentive. The groups are defined as follows: devices only (Group D), devices and mobile app (Group D+A), and devices and mobile app with a financial incentive (Group D+A+F). Group D received only the devices (i.e., the activity tracker and scale). Group D+A received the devices as well as a smartphone application developed by the study team. Group D+A+F received the devices, the study smartphone application, and a financial incentive based on the participant’s adherence, which maxed out at $150 and was paid at the completion of the monitoring period. Over the phone, all participants were taught how to use the devices, and those in Groups D+A and D+A+F were also taught how to access the smartphone application and navigate all features within the application. Groups were also measured using standard surveys (described below).

### Study Devices and Mobile App

#### Fitbit Charge 4 (FC4) and Fitbit Charge 5 (FC5)

The Fitbit Charge 4 (FC4) and Fitbit Charge 5 (FC5) were used in all groups to provide daily feedback on a variety of parameters, such as physical activity. The FC4 (Fitbit Inc.) is a commercially available wrist□worn device with a 3-axial accelerometer, an altimeter, and an optical heart rate tracker. Similarly, the FC5 (Fitbit Inc.) consists of a 3-axial accelerometer, vibration motor, multipurpose electrical sensors, and an optical heart-rate tracker. Using these components, the FC4 and FC5 track, record, and deliver real-time information on step count, heart rate, sleep, and active minutes. Patients were directed to wear the Fitbit device on their wrists at all times, except when charging the device. Patients were also instructed on how to use the Fitbit smartphone application. The device’s battery life lasts between three and five days, with a charging time of approximately one hour.

#### BodyTrace Scale

Participants were provided with a BodyTrace scale (BodyTrace Inc., New York, New York) for daily weighing. The BodyTrace scale includes a cellular modem with a factory-installed SIM card. The scale arrives ready for use with four AA batteries and requires no user configuration. Weight data is uploaded to a cloud-based database via the cellular modem. From this database, researchers can directly download the data.

#### myHeartCare Mobile Application

For this study, the research team developed *myHeartCare*, a cross-platform (iOS or Android) mobile app capable of linking to the Fitbit tracker and BodyTrace scale. Groups D+A and D+A+F, the mobile app users, downloaded this app and were instructed on navigating its features. The app is divided into four sections: 1) adherence statistics, 2) surveys, 3) rewards, and 4) social. A daily “To-Do” list on the statistics page prompts patients to sync their Fitbit to their smartphone, weigh themselves, and complete a daily survey via the app, which used the same three questions as described above regarding diet, medication, and HF symptoms. At 9 AM each day participants received a push notification asking them to sync their Fitbit and take their daily survey. On the statistics page, participants can also track their weight over time and identify fluctuations by visualizing the recorded data from the BodyTrace scale as a time plot. The survey page enables participants to access their daily surveys and the rewards page allows them to monitor their study progress. For adhering to their daily tasks (syncing Fitbit, weight, survey), participants in Group D+A were able to earn and track points. Those in Group D+A+F were also able to accumulate and track points with the addition that points were tied to financial earnings, which were displayed to the user and delivered via a cash gift card at the end of the 180-day study period. Additionally, the social page allowed users to invite friends and family to view their adherence history and send encouraging messages.

### Baseline Survey

Those who consented to participate in the study were administered a baseline survey consisting of questions relating to demographic information and two institutional review board-approved questionnaires—the Minnesota Living with Heart Failure Questionnaire (MLHFQ) (Bilbao et al., 2016) and Patient-Reported Outcomes Measurement Information System (PROMIS) Global Health (Cella et al., n.d.). These questionnaires were used to measure the HF patients’ health-related quality of life through patient-reported outcomes using a 5-point Likert scale. Following this survey, patients were officially consented into the study and randomized to one of the three study groups.

### Follow-Up Survey

After 180 days of monitoring, participants were contacted over the phone and administered a follow-up survey. The survey consisted of the same two questionnaires administered in the baseline survey—the MLHFQ and PROMIS Global Health. In addition to these questionnaires, participants were asked to rate their experiences with the Fitbit and the BodyTrace scale. The survey assessed factors of these devices, such as helpfulness, ease of incorporation into daily life, and whether each device helped the participant adhere to their care plan. Participants were then offered an option for an additional 180 days of Fitbit and BodyTrace scale data collection, which did not require completion of the daily survey.

### Data Collection and Analysis

The study team configured a server to connect to vendor APIs that were used to pull participant data and update the myHeartCare app in real-time when a task had been completed for subjects in the D+A and D+A+F groups. The server also received the responses to the daily questionaire for the D+A and D+A+F groups. Three metrics were used to define adherence: if a subject 1) synced their Fitbit, 2) checked their weight, and 3) completed the daily survey. Adherence for each task is computed as the fraction of days the task was completed (e.g. Fitbit synced) divided by the total number of days in the time period of analysis (e.g., 30 days). The three groups were compared across the three measurements of adherence using Kruskal-Wallis tests and Mann-Whitney U tests to make pairwise comparisons.

## Results

111 participants completed the 180 study between July 2021 and April 2023. The demographics of the participants who completed the study are shown in Table 1. Our results demonstrate that participants within groups D+A and D+A+F had a higher adherence rate over time for the three measured tasks compared to Group D. Moreover, there was a greater decline in Group D’s adherence rates as their participation in the study progressed relative to the groups which utilized the mobile app. Figure 2 displays the average adherence of each group to the three measured tasks for each day of participation in the study and Figure 3 shows box plots of subject adherence over 30 day and 180 day periods.

**Figure 2.**
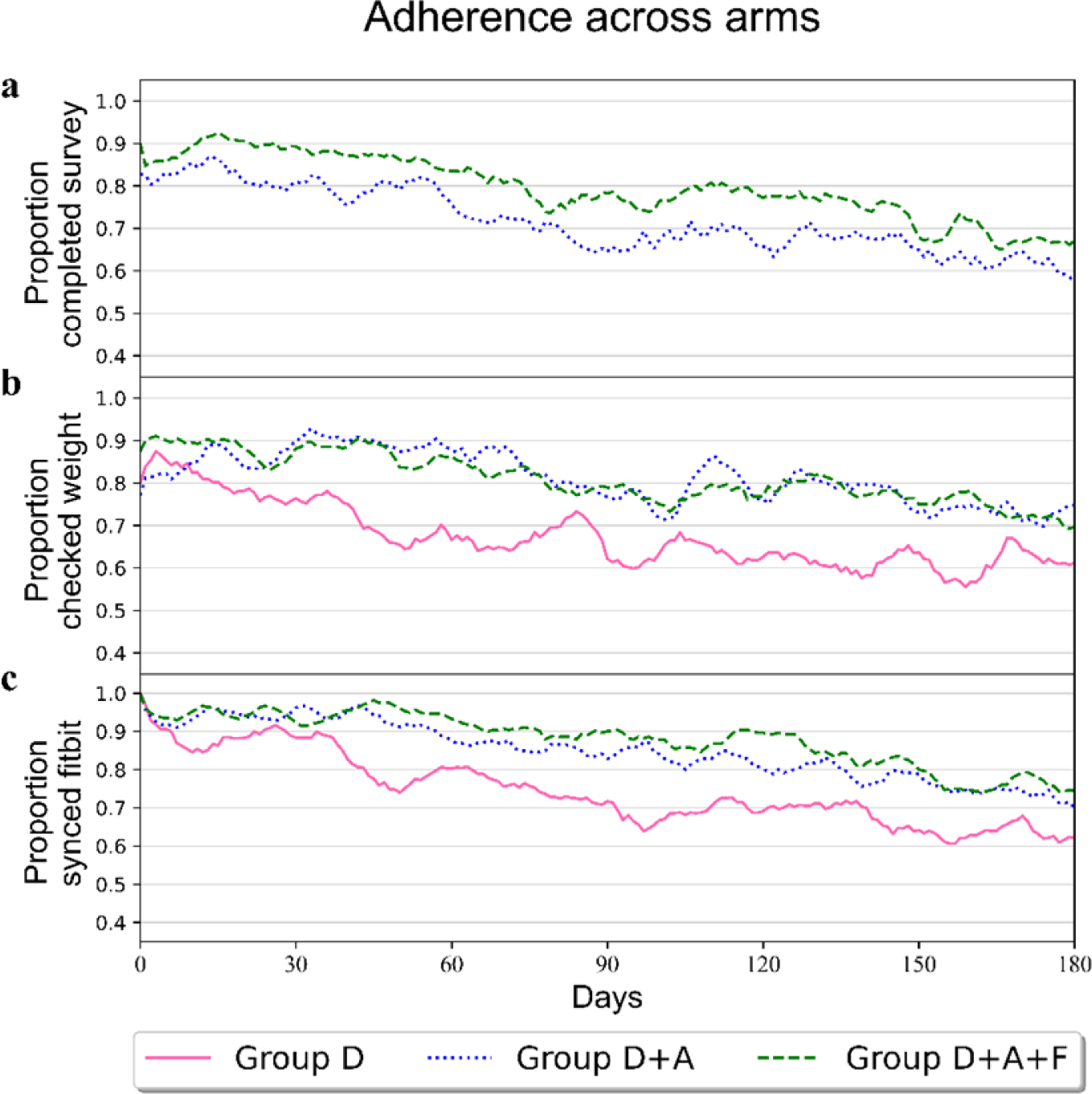
Plots of adherence rates for the three primary tasks completed by study participants, classified by intervention group. Group D is classified with a solid pink line, Group D+A is classified with a dotted blue line, and Group D+A+F is classified with a dotted green line.

**Figure 3.**
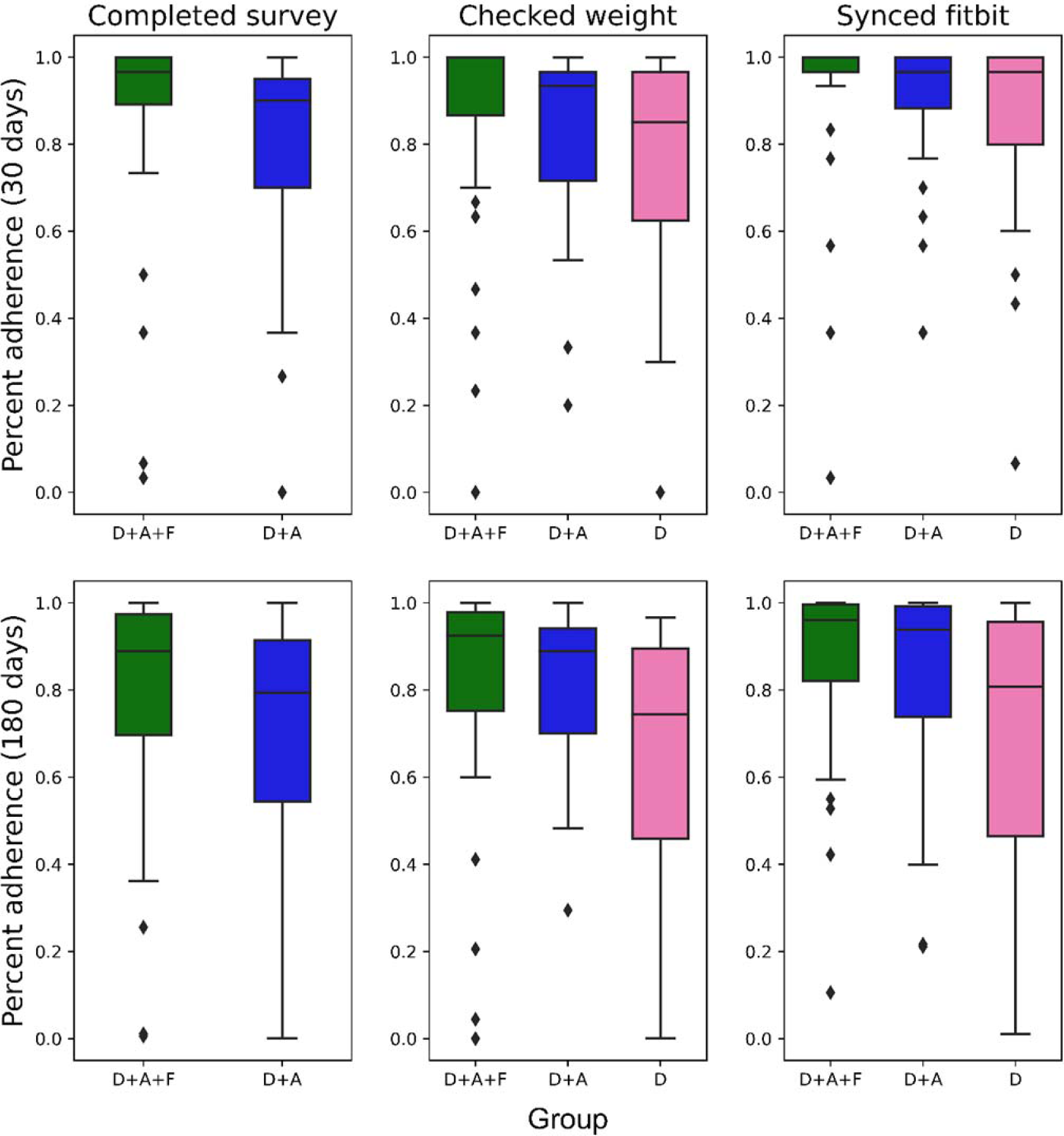
Box plots of device adherence and mobile app survey completion (D+A and D+A+F groups only) for the different groups at 30 and 180 days. Unit of analysis is a subject’s fraction of days the task was completed divided by the time period. Two subjects did not have cellular service at their home, which prevented the weight scale from syncing and thus resulted in 0% adherence for the duration of the study.

**Table 1.** Demographics of 111 participants who completed the study, organized by category.

| Category | Sub-Category | Frequency (N) | Percent (%) |
| --- | --- | --- | --- |
| Gender | Male | 82 | 73.87% |
|  | Female | 29 | 26.13% |
|  | Non-binary/third gender | 0 | 0.00% |
|  | Prefer not to say | 0 | 0.00% |
| Race | White | 83 | 74.77% |
|  | Black/African-American | 15 | 13.51% |
|  | Asian | 10 | 9.01% |
|  | American Indian/Alaskan | 3 | 2.70% |
|  | Native Hawaiian/Pacific Islander | 0 | 0.00% |
|  | Don't know/Refuse | 5 | 4.50% |
| Age | 50-60 | 33 | 26.73% |
|  | 61-70 | 41 | 36.94% |
|  | 71-80 | 37 | 33.33% |
| Education | High School | 7 | 6.31% |
|  | Some College/Associate Degree/Trade School | 27 | 24.32% |
|  | Bachelor's Degree | 25 | 22.52% |
|  | Master's Degree or above | 52 | 46.85% |
|  | Don't know/Refuse | 0 | 0.00% |
| Income | \$0-\$25,000 | 8 | 7.21% |
| | \$25,001-\$50,000 | 12 | 10.81% |
| | \$50,001-\$75,000 | 8 | 7.21% |
| | \$75,001 or more | 77 | 69.37% |
|  | Don't know/Refuse | 6 | 5.41% |
| Hispanic/Latino | No | 101 | 91.00% |
|  | Yes | 10 | 9.00% |
|  | Don't know/Refuse | 0 | 0.00% |
| Ischemic/Non-ischemic | Ischemic | 58 | 52.25% |
|  | Non-ischemic | 43 | 38.74% |
|  | Unknown | 10 | 9.01% |
| NYHA Class | 1 | 13 | 11.71% |
|  | 2 | 43 | 38.74% |
|  | 3 | 21 | 18.92% |
|  | 4 | 2 | 1.80% |
|  | 1 to 2 | 3 | 2.70% |
|  | 2 to 3 | 9 | 8.10% |
|  | 3 to 4 | 2 | 1.80% |
|  | Unknown | 18 | 16.22% |
| LVEF Median | <10 | 1 | 0.9% |
|  | 15-20 | 2 | 1.80% |
|  | <20 | 1 | 0.9% |
|  | 20-25 | 6 | 5.41% |
|  | 25-30 | 6 | 5.41% |
|  | 30-35 | 16 | 14.41% |
|  | 35-40 | 11 | 9.91% |
|  | <40 | 1 | 0.9% |
|  | 40-45 | 8 | 7.21% |
|  | 45-50 | 10 | 9.01% |
|  | 50-55 | 12 | 10.81% |
|  | 55-60 | 9 | 8.11% |
|  | 60-65 | 8 | 7.21% |
|  | 65-70 | 2 | 1.80% |
|  | 70-75 | 3 | 2.70% |
|  | Unknown | 15 | 13.51% |

### Fitbit Syncing

Regarding Fitbit syncing, those in group D+A+F had an adherence rate of 97.5% after the first 30 days in the study and an adherence rate of 95% at the end of the 180-day period. Those in group D+A had an adherence rate of 100% after the first 30 days and 91.4% at the end of the study for syncing their Fitbit. Group D participants had an adherence rate of 97.2% after the first 30 days and an adherence rate of 72.2% at the end of the study for syncing their Fitbit. A significant difference in adherence (p < 0.05) was found between Group D and Group D+A+F during the first 30 (p=0.0097), 60 (p=.0028), 90 (p=.0051), 120 (p=.0081), 150 (p=0.0061), and 180 (p=0.0101) days of the study.

### Self-Weighing

For self-weighing, Group D+A+F had an adherence rate of 92.5% after the first 30 days and 87.5% at the end of the study. Group D+A had an adherence rate of 94.3% after the first 30 days and ended with the same adherence rate for weighing themselves at the 180-day time point. For Group D, the adherence rate was 94.4% after 30 days and 69.4% at the 180-day period of the study ending. Statistical analysis revealed a significant difference between Group D and Group D+A+F during the first 30 (p=0.0005), 60 (p=0.0001), 90 (p=0.0001), 120 (0.0006), 150 (p=0.0010), and 180 (p=0.0016) days of the study. There was a significant difference between Group D+A and Group D+A+F only for the first 60 (p=0.031) days of the study. A significant difference was found between Group D and Group D+A for the first 60 (p=0.0166), 90 (p=0.0119), 120 (p=0.0106), 150 (p=0.0138), and 180 (p=0.0201) days of the study.

### Survey Taking

Participants in Group D did not have the option of taking the daily survey as they did not have the mobile app. Group D+A+F had an adherence rate of 95% at 30 days and 87.5% at 180 days. Group D+A had adherence rates of 88.6% after the first 30 days and 82.9% at the end of the study. Statistical analysis indicated only a significant difference between Group D+A and Group D+A+F for the first 30 (p=0.0391) and 60 (p=0.0347) days of the study.

## Conclusions

Research investigating the value of remote monitoring in HF patients has generally found adherence to be a critical barrier to effective implementation. Another study of HF patients used a wireless transmission pod, a scale, and a blood pressure and heart rate monitor to capture data remotely (Ong et al., 2016). At 180 days, the investigators found 68% adherence to telephone coaching, but only 51.7% adherence to telemonitoring. A previous study conducted by our team employed Fitbits in the remote monitoring of ischemic heart disease patients and observed an adherence rate to daily syncing of 72% at the 90-day time point (Speier et al., 2018). This result is similar to Group D’s adherence rate in the current study. In contrast, the adherence rates at 90 days for Groups D+A and D+A+F were 82.86% and 90%, respectively. Notably, the inclusion of a financial incentive (Group D+A+F) drove adherence significantly higher than the baseline group with no app (Group D), and even Group D+A had significantly higher adherence than Group D for weight monitoring. These findings support the use of mobile apps to boost adherence to telemonitoring.

Despite higher adherence relative to previous work, we still observed adherence decay over time. Future modifications to our study protocol could incorporate an incentive at regular time intervals to sustain long-term adherence, such as device upgrades and more frequent financial reward distributions rather than waiting until the end of the study. Additionally, real-time responses could be solicited via the app to participants who begin falling in adherence, providing new insights and possibly ways to improve engagement.

Finally, given that the app, tracker, and scale send data in real time, there is an opportunity for scalable monitoring of populations using machine learning approaches that could search for patients at risk of exacerbation. These algorithms would require larger datasets with which to train, as well as targets defined from clinical review or patient follow-up (e.g., urgent care visits). The inclusion of additional data points, such as demographics and co-morbidities, would likely improve algorithm performance. Similarly, methods for tracking symptoms, diet, and medication adherence more quantitatively and precisely would be important indicators for adverse future events.

## Data Availability

The data used in this study is available upon request.

## Notes

### Competing Interest Statement

The authors have declared no competing interest.

### Funding Statement

This work was support by the NIH/NHLBI under grant number R01HL141773

### Author Declarations

This prospective study included patients with heart failure at the University of California, Los Angeles (UCLA) Medical Center. The study was approved by the UCLA Institutional Review Board (IRB #19-000399).

